# Disentangling post-vaccination symptoms from early COVID-19

**DOI:** 10.1101/2021.07.21.21260906

**Authors:** Liane S. Canas, Marc F. Österdahl, Jie Deng, Christina Hu, Somesh Selvachandran, Lorenzo Polidori, Anna May, Erika Molteni, Benjamin Murray, Liyuan Chen, Eric Kerfoot, Kerstin Klaser, Michela Antonelli, Alexander Hammers, Tim Spector, Sebastien Ourselin, Claire Steves, Carole H. Sudre, Marc Modat, Emma L. Duncan

## Abstract

**Background:** Identifying and testing individuals likely to have SARS-CoV-2 is critical for infection control, including post-vaccination. Vaccination is a major public health strategy to reduce SARS-CoV-2 infection globally. Some individuals experience systemic symptoms post-vaccination, which overlap with COVID-19 symptoms. This study compared early post-vaccination symptoms in individuals who subsequently tested positive or negative for SARS-CoV-2, using data from the COVID Symptom Study (CSS) app.

**Design:** We conducted a prospective observational study in UK CSS participants who were asymptomatic when vaccinated with Pfizer-BioNTech mRNA vaccine (BNT162b2) or Oxford-AstraZeneca adenovirus-vectored vaccine (ChAdOx1 nCoV-19) between 8 December 2020 and 17 May 2021, who subsequently reported symptoms within seven days (other than local symptoms at injection site) and were tested for SARS-CoV-2, aiming to differentiate vaccination side-effects *per se* from superimposed SARS-CoV-2 infection. The post-vaccination symptoms and SARS-CoV-2 test results were contemporaneously logged by participants. Demographic and clinical information (including comorbidities) were also recorded. Symptom profiles in individuals testing positive were compared with a 1:1 matched population testing negative, including using machine learning and multiple models including UK testing criteria.

**Findings:** Differentiating post-vaccination side-effects alone from early COVID-19 was challenging, with a sensitivity in identification of individuals testing positive of 0.6 at best. A majority of these individuals did not have fever, persistent cough, or anosmia/dysosmia, requisite symptoms for accessing UK testing; and many only had systemic symptoms commonly seen post-vaccination in individuals negative for SARS-CoV-2 (headache, myalgia, and fatigue).

**Interpretation:** Post-vaccination side-effects per se cannot be differentiated from COVID-19 with clinical robustness, either using symptom profiles or machine-derived models. Individuals presenting with systemic symptoms post-vaccination should be tested for SARS-CoV-2, to prevent community spread.

**Funding:** Zoe Limited, UK Government Department of Health and Social Care, Wellcome Trust, UK Engineering and Physical Sciences Research Council, UK National Institute for Health Research, UK Medical Research Council and British Heart Foundation, Alzheimer’s Society, Chronic Disease Research Foundation, Massachusetts Consortium on Pathogen Readiness (MassCPR).

**Research in context:** *Evidence before this study:* There are now multiple surveillance platforms internationally interrogating COVID-19 and/or post-vaccination side-effects. We designed a study to examine for differences between vaccination side-effects and early symptoms of COVID-19. We searched PubMed for peer-reviewed articles published between 1 January 2020 and 21 June 2021, using keywords: “COVID-19” AND “Vaccination” AND (“mobile application” OR “web tool” OR “digital survey” OR “early detection” OR “Self-reported symptoms” OR “side-effects”). Of 185 results, 25 studies attempted to differentiate symptoms of COVID-19 vs. post-vaccination side-effects; however, none used artificial intelligence (AI) technologies (“machine learning”) coupled with real-time data collection that also included comprehensive and systematic symptom assessment. Additionally, none of these studies attempt to discriminate the early signs of infection from side-effects of vaccination (specifically here: Pfizer-BioNTech mRNA vaccine (BNT162b2) and Oxford-AstraZeneca adenovirus-vectored vaccine (ChAdOx1 nCoV-19)). Further, none of these studies sought to provide comparisons with current testing criteria used by healthcare services.

*Added value of this study:* This study, in a uniquely large community-based cohort, uses prospective data capture in a novel effort to identify individuals with COVID-19 in the immediate post-vaccination period. Our results show that early symptoms of SARS-CoV-2 cannot be differentiated from vaccination side-effects robustly. Thus, post-vaccination systemic symptoms should not be ignored, and testing should be considered to prevent COVID-19 dissemination by vaccinated individuals.

*Implications of all the available evidence:* Our study demonstrates the critical importance of testing symptomatic individuals - even if vaccinated – to ensure early detection of SARS-CoV-2 infection, helping to prevent future pandemic waves in the UK and elsewhere.

## Introduction

The havoc wrought by SARS-CoV-2 is unprecedented in living memory, with >184 million cases of COVID-19 world-wide and >4.0 million deaths by 8 July 2021.^1,2^ Extraordinary efforts directed towards rapid vaccine development meant that by late 2020 the UK Medicines and Healthcare products Regulatory Agency had authorized three vaccines: Pfizer-BioNTech mRNA (BNT162b2)^3,4^ Oxford-AstraZeneca adenovirus-vectored^5–7^ and Moderna mRNA (mRNA-1273)^8,9^ A fourth vaccine (Janssen adenovirus-vectored Ad26.COV2.S) was authorised on 28 May 2021.^10^ Vaccination with BNT162b2 (herein, PB) and ChAdOx1 nCoV-19 (herein, O-AZ) started in the UK on 8 December 2020^11^ and 4 January 2021^12^ respectively, during which time the UK was experiencing its third pandemic wave with widespread community transmission (peak UK positive specimens reported on 29 December 2020^13^). Since then, and in the context of social distancing and stay-at-home directives, new infections, hospitalisations, and deaths from SARS-CoV-2 have fallen rapidly. ^1,2,14^

Local and systemic reactions have been observed after all vaccines for SARS-CoV-2. Considering the two vaccines used predominantly in the UK to date (O-AZ and PB), local reactions were common during their pivotal trials (76% of younger (<55 years) O-AZ recipients reported tenderness;^5,6^ 83% of younger PB recipients reported pain).^4^ Systemic reactions were also common and included fatigue (O-AZ 76%; PB 59%), headache (O-AZ 65%; PB 52%), and fever (O-AZ 24%; PB 16%).^4–6^ Observational data from the COVID Symptom Study (CSS)^15^ also showed high incidence of local (62%) and systemic (26%) effects.^16^ The most serious side-effect to date, observed after both O-AZ and Janssen vaccines, is vaccine-induced immune thrombotic thrombocytopenia (VITT) associated with anti-PF4 antibody production.^17,18^ As of 9 June 2021, with 69,743,980 vaccinations administered in the UK (>45 million O-AZ, including first and second doses), 395 cases of VITT have been reported, with 70 deaths.^19^ Saliently, most vaccine-related side-effects (including VITT) are more common in younger individuals, whereas COVID-19 clinical severity increases with age.^4–6,16^

Prevention of SARS-CoV-2 dissemination requires rapid recognition followed by quarantining of infected individuals (along with appropriate health care). However, there is overlap between symptoms from COVID-19^20,21^ and early post-vaccination systemic symptoms.^4–6,16^ Moreover, immunity to SARS-CoV-2 does not occur immediately post-vaccination,^22^ with functional protection from approximately day 12.^23^ Quarantining and testing every individual with systemic symptoms early post-vaccination would be onerous, expensive, and labour-intensive – but given the impact of viral outbreaks might be unavoidable if SARS-CoV-2 infection cannot be excluded robustly.^20,21^

Here we aim to determine whether symptom profiles can be used to differentiate individuals with systemic side-effects of vaccination alone from individuals with superimposed SARS-COV-2 infection.

## Methods

### Study design and Participants

Data were acquired prospectively from the CSS, using a mobile health application launched by ZOE Limited and King’s College London in March 2020 (app details and development given in Supplementary Methods).^15,21^ Briefly, individuals are asked daily to log their health status, health care access, SARS-CoV-2 testing and results, and vaccination details, with direct questions about symptoms associated with COVID-19 (Supplementary Table S1).^14,15,21^ Symptomatic individuals are prompted to undergo testing, either through standard care or through test request from ZOE/CSS.^24^

Data were acquired from UK participants aged 16-90 years, between 8 December 2020 (UK vaccination start date) and 17 May 2021, who were asymptomatic when vaccinated with PB or O-AZ (first or second dose), and subsequently reported: a) at least one predefined symptom (Supplementary Table S1) within seven days post-vaccination, and b) a SARS-CoV-2 test result (rtPCR or lateral flow antigen test [LFAT]) within ten days post-vaccination. The seven-day cut-off for symptom presentation was informed by: a) serial interval for COVID-19 (mean, 5.2 days^25)^; b) incubation period for SARS-CoV-2 (mean, 5.8 days^25,26^); c) the timeline for acute post-vaccination side-effects in both pivotal trials (one-week^4–6,8,10^) and d) reported real-world experience of post-vaccination symptoms (peak prevalence day 1 post-vaccination; mean duration one day^16^). The ten-day cut-off for testing allowed three days’ delay in accessing testing.^27^

Early results indicated a large imbalance in numbers of individuals testing positive vs. negative post-vaccination, sufficient to bias analysis.^28^ A 1:1 population from the negative cohort (matching age, BMI, gender, occupation, week of testing, and comorbidities) was selected based on minimisation of Euclidean distance between positive and negative subjects considering these features, enabling a fair comparison between groups of equal size.^29,30^ However, to ensure robustness, analyses were repeated using: a) a one-hundred bootstrapping scheme selecting from the negative population; and b) the entire negative population.

Individual symptoms (here, a symptom reported at any time within seven days post-vaccination, irrespective of duration) were compared between vaccinated individuals testing positive or negative for SARS-CoV-2, using Chi-squared tests per symptom. Duration of individual symptoms was calculated as days from first report of that symptom, until asymptomatic and/or seven days post-vaccination, noting that duration beyond seven days was not considered; however, as number of individuals experiencing each symptom were low in both groups, no statistical comparison was made. Symptom burden, defined as total symptom count per person [irrespective of symptom duration] were compared between groups using Mann-Whitney-U tests. We also considered symptom manifestation across the week post-vaccination, by dynamic profiling for each symptom (symptom frequency). Distribution of symptom duration was assessed using Mann-Whitney-U. Correlation of individual symptoms within both positive and negative individuals was assessed by computing a Spearman-rank correlation test. Local symptoms due to vaccination *per se* (Supplementary Table S2) were excluded from analysis as unlikely to be indicative of, or influenced by, SARS-CoV-2 infection.

Machine learning was used to determine if post-vaccination symptoms *per se* could be separated from superimposed SARS-CoV-2 infection (including symptom combination, and cumulative symptom burden).^16,21^ We trained a set of binary classifiers to identify SARS-CoV-2-positive individuals. Models included random forest, logistic regression, and Bayesian mixed-effect models, exploiting their varying properties (Supplementary Table S3) to improve reliability of results. We also considered whether symptoms were after first or second vaccination and the impact of vaccination order, by including vaccination ordering as a covariate. Models were trained on data without stratifying by vaccine type, due to small sample sizes. We also did not discriminate between type of SARS-CoV-2 testing (PCR vs. LFAT), or mode of testing access (NHS vs. ZOE-request), either in model development or other analyses.

We sought to reduce bias from assessing high numbers of individual symptoms by performing symptom-clustering using K-means. ^31^ However, a relevant/accurate number of clusters was not evident from the silhouette plot and entropy (data not shown); thus further analyses using machine clustering were not pursued. Symptoms were clustered manually into clinical groupings (reviewed by ELD, MO, TS, AH, CJS) (Supplementary Table S4), and analysed using the above models similarly. Lastly, clustering based on having at least one of the four symptoms required for accessing NHS testing during the timing of this study (viz., presence or absence of fever, persistent cough, anosmia and/or dysosmia)^24^ were assessed.

Data were split into training and validation sets for random forest, logistic regression, and Bayesian mixed-effect models. Five folds were used on the training set, composed of 80% of the initial dataset randomly selected, to train the models in different subsamples of the population. The remaining 20% were then used to assess the performance of models, evaluating sensitivity, specificity, and balanced accuracy. The class ratio was maintained in both training and testing sets. For fair evaluation, both clinical clustering and categorisation using the NHS criteria were assessed on 20% of the data of each fold.

### Role of the funding source

ZOE Limited developed the app for data collection as a not-for-profit endeavour. The funder had no role in study design, data analysis, data interpretation, or influence on report content.

### Ethical approval

The app and CSS were approved in the UK by KCL’s ethics committee (REMAS no. 18210, review reference LRS-19/20–18210). All app users provided informed consent for use of their data for COVID-19 research.

## Results

Figure 1 shows the flow chart for this study. Overall, 1,072,313 UK CSS app users were vaccinated with either O-AZ or PB (O-AZ: 713,651; PB: 358,662). Of these, 362,770 (33.8% (O-AZ: 264,587; PB: 98,183)) reported at least one symptom early post-vaccination, with SARS-CoV-2 testing in 14,842 (4.09%) of these individuals (O-AZ: 10,765; PB: 4,077). A positive test was reported by 150 (1.01%) individuals (O-AZ: 73/10,765 (0.68%); PB: 77/4077 (1.89%)).

**Figure 1.**
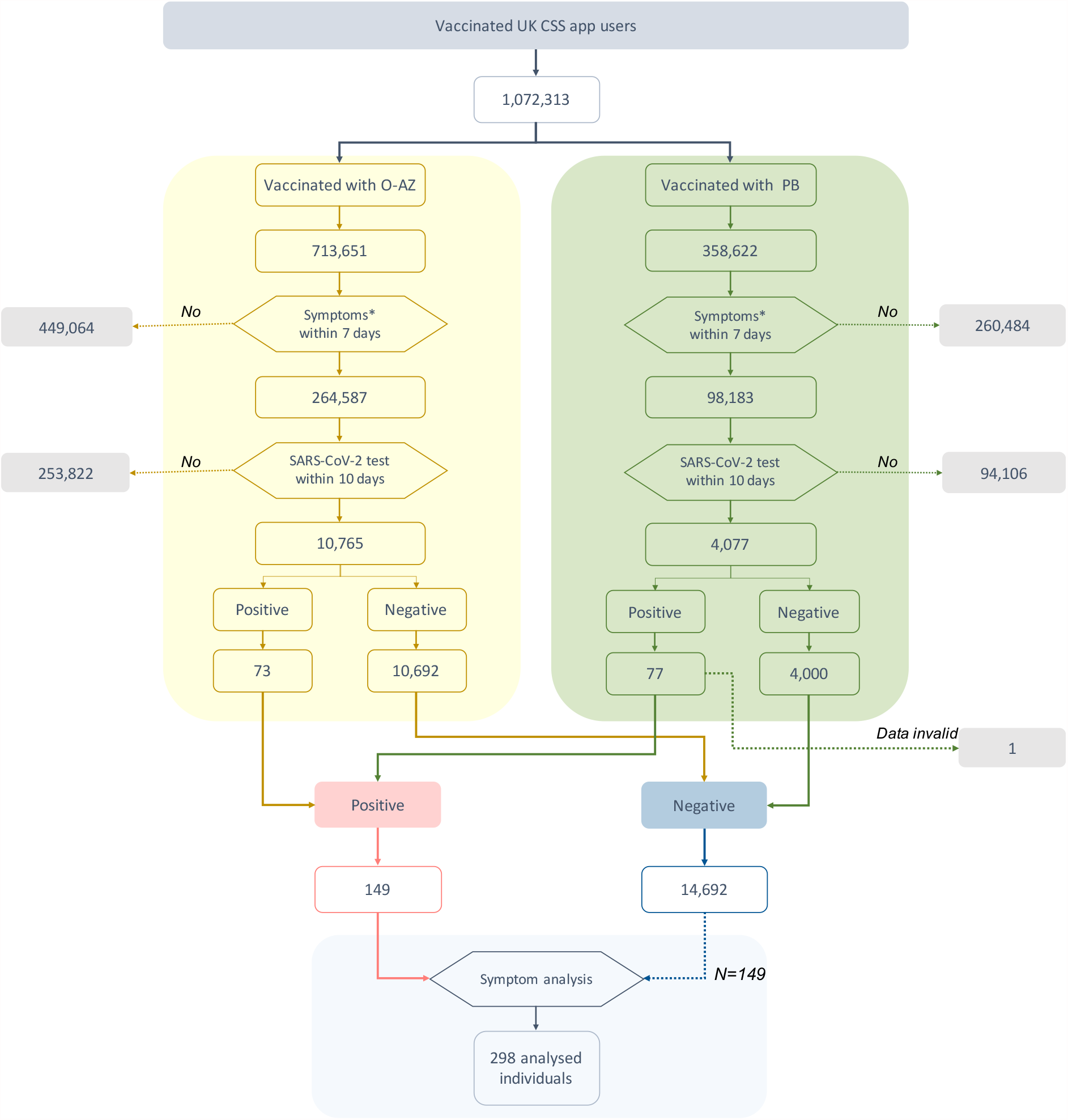
Flowchart of individuals included in this study. Symptoms^*^ within 7 days excluded local symptoms related to injection site. SARS-CoV-2 test included both rtPCR and LFAT. Positive and negative refers to self-logged test results.

Within the tested group, 3,525/14,842 (23.75%) reported at least one requisite symptom fulfilling UK testing criteria ^24^; 62 (1.76%) tested positive. Conversely, 11,317 tested individuals did not report any requisite symptom, of whom 88 (0.78%) tested positive. Individuals with requisite symptoms were more likely to test positive than those without (p-value<0.0001); none-the-less, the majority (88 of 150, 59%) who tested positive did not meet current UK testing criteria.

For further analyses, one positive individual (vaccinated with PB) was excluded due to invalid data entry (invalid BMI), leaving 149 symptomatic positively-tested individuals. Table 1 describes the positive and matched negative cohorts.

**Table 1.** Demographic information of vaccinated individuals testing positive or negative for SARS-CoV-2 infection. Data are presented as median value [IQR] for age and BMI; and numbers (percentages) for other values. *BMI: Body Mass Index. IQR: Inter-Quartile Range*.

|  | Vaccinated Cohort |  |  |  |  |  |
| --- | --- | --- | --- | --- | --- | --- |
|  | Positive testing for SARS-CoV-2 infection |  |  | Negative testing for SARS-CoV-2 infection |  |  |
|  | O-AZ | PB | Full cohort | O-AZ | PB | Full cohort |
| Number | 73 | 76 | 149 | 73 | 76 | 149 |
| Males (%) | 27 (37.0) | 21 (27.6) | 48 (32.2) | 22 (30.1) | 19 (25.0) | 45 (30.2) |
| Age, years (median [IQR]) | 62.0 [50.0; 71.0] | 59.0 [50.0; 67.5] | 61.0 [50.0; 70.0] | 65.0 [54.0; 69.0] | 64.0 [52.0; 71.3] | 63.0 [52.0; 70.0] |
| BMI (median [IQR]) | 25.0 [22.7; 28.0] | 26.1 [23.5; 29.3] | 25.4 [23.4; 29.2] | 24.9 [23.0; 28.1] | 25.5 [23.3; 27.0] | 25.2 [23.3; 28.6] |
| Lung disease (%) | 8 (11.0) | 9 (11.8) | 17 (11.4) | 8 (11.0) | 14 (18.4) | 20 (13.4) |
| Kidney Disease (%) | 0 (0.0) | 1 (1.3) | 1 (0.7) | 1 (1.4) | 0 (0.0) | 1 (0.7) |
| Diabetes (%) | 4 (5.5) | 5 (6.6) | 9 (6.0) | 4 (5.5) | 3 (3.9) | 9 (6.0) |
| Heart Disease (%) | 7 (9.6) | 4 (5.3) | 11 (7.4) | 2 (2.7) | 4 (5.3) | 5 (3.4) |
| Cancer (%) | 0 (0.0) | 4 (5.3) | 4 (2.7) | 4 (5.5) | 3 (3.9) | 6 (4.0) |
| Healthcare workers (%) | 0 (0.0) | 10 (13.2) | 10 (6.7) | 0 (0.0) | 7 (9.2) | 2 (1.3) |
| Visit to hospital (%) | 1 (1.4) | 1 (1.3) | 2 (1.3) | 1 (1.4) | 0 (0.0) | 2 (1.3) |

Four symptomatic individuals who tested positive did so within 10 days of their second vaccination; their symptoms after first vaccination were thus disregarded. As the matched negative control cohort (N=149) included matching for vaccination order, the controls also included four individuals reporting symptoms after second vaccination. Given the small sample size, all 149 subjects in each of the positive and negative groups were included in model training, which included regressing data from vaccination order. However, as we could not conduct a fair statistical analysis of symptoms after first vs. second vaccination for symptom profiling (given the very small numbers of infected individuals presenting after second vaccination) and as post-vaccination symptoms vary after first vs. second dose,^16,32^ we present the symptom profiles after first vaccination (N=145) in the main text with data from the entire cohort (N=149) in the Supplementary Materials (Supplementary Table S5).

Individual symptom prevalence after first vaccination is shown in Figure 2 and Supplementary Table S5. Although some symptoms were more common in individuals testing positive vs. negative (sore throat (p-value = 0.0187), sneezing (p value = 0.0474) and persistent cough (p-value = 0.0396)), others were more common in the negative group (palpitations (p-value = 0.0284). The numbers of individuals reporting each symptom were small (e.g., sore throat, n=17; persistent cough, n =12) (Supplementary Table S5).

**Figure 2.**
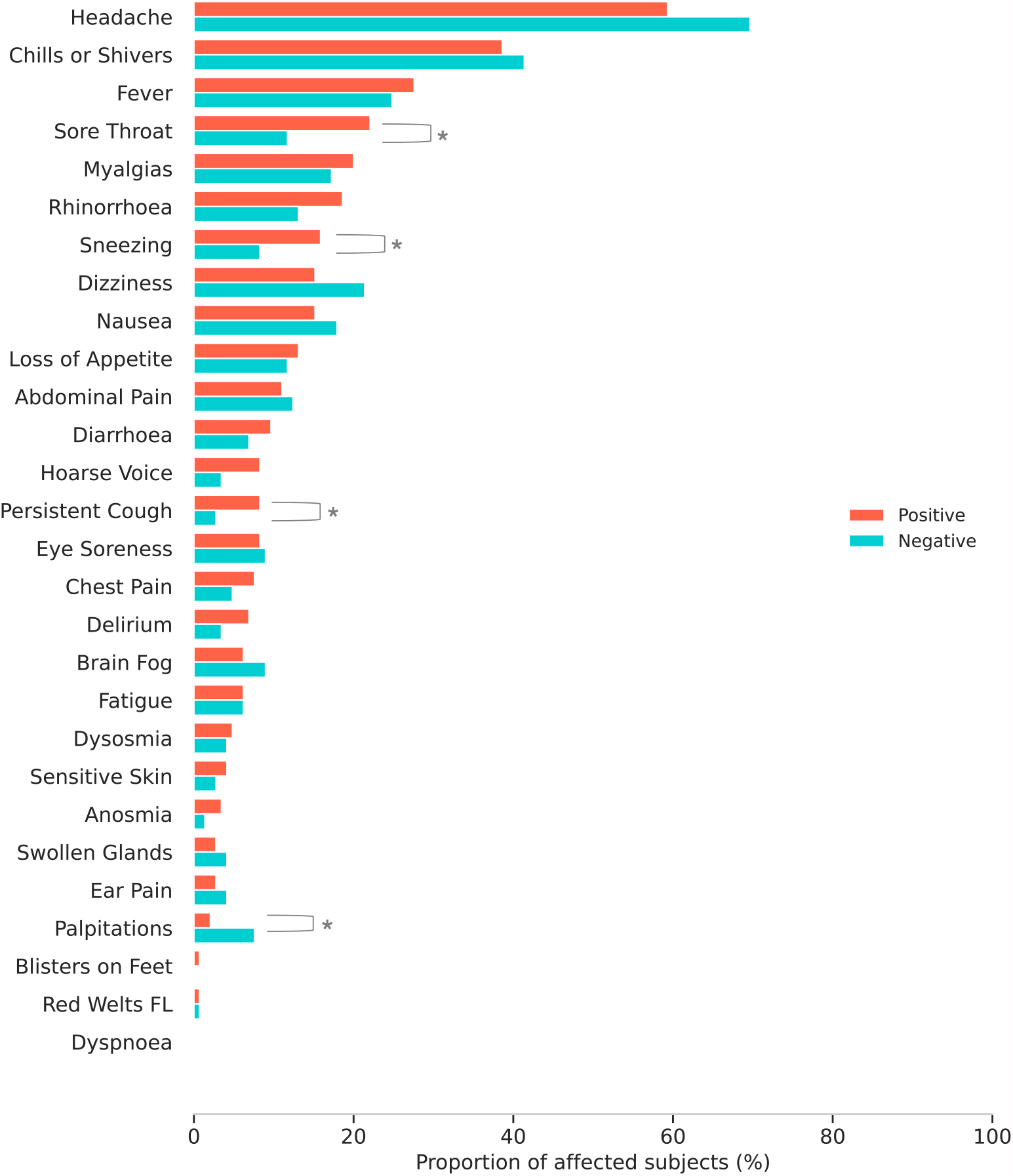
Profiles of illness in symptomatic individuals early post-vaccination, comparing symptom prevalence (symptom reported at any time during first week) in positive vs. negative cases (1:1 matched population; N=145 for each). ^*^p < 0.05 ^**^p < 0.01.

Median day of onset for any symptom post-vaccination was Day 1 in both groups (noting all individuals were asymptomatic when vaccinated), with highest symptom burden on Day 3, again in both groups (Figure 3; and Supplementary Tables S6 and S7). There was no difference in symptom burden between individuals testing positive or negative (median: 12 in positive group, 10.5 in negative group, Mann-Whitney test p-value = 0.22).

**Figure 3.**
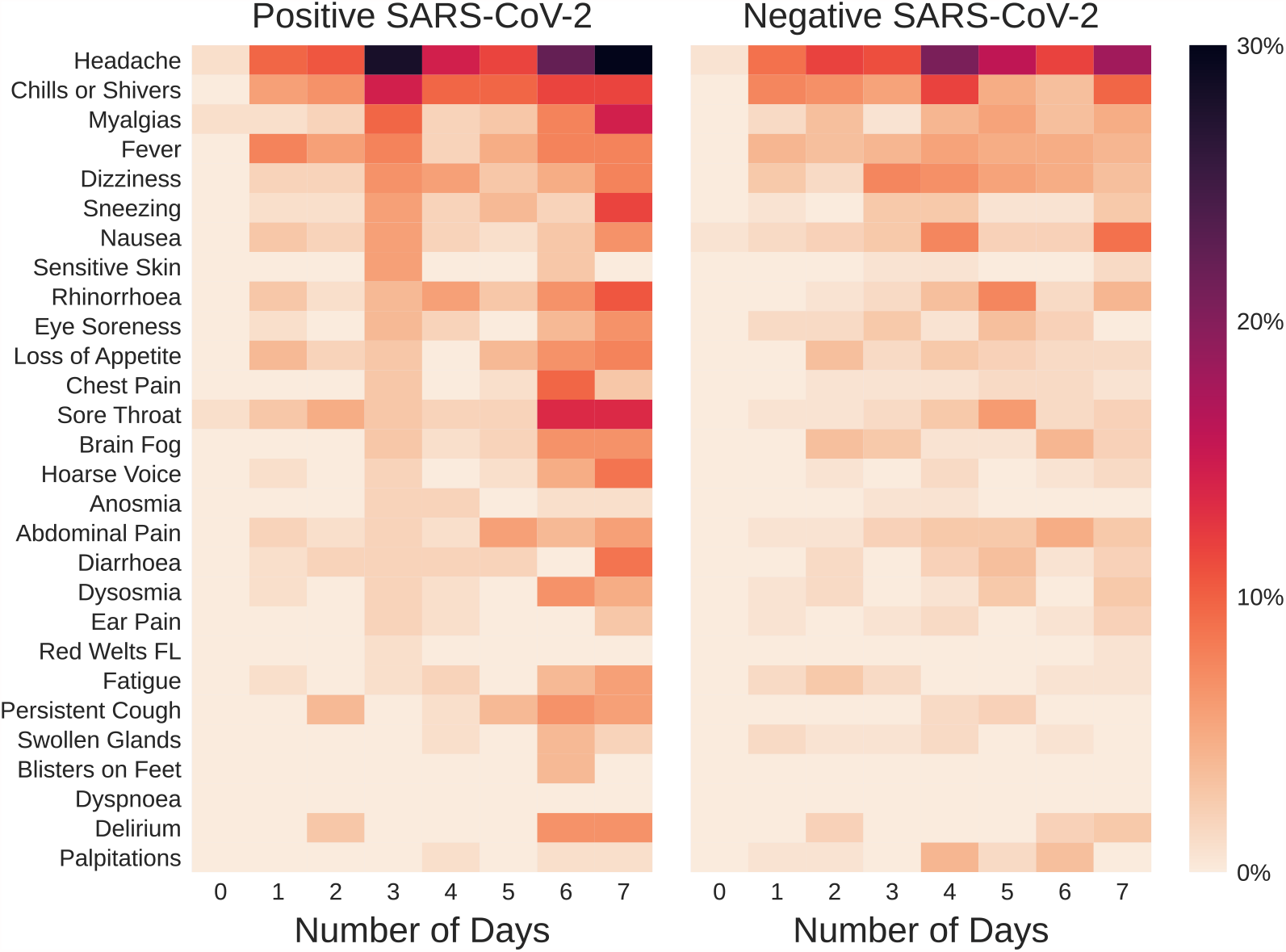
Symptom prevalence and distribution during the first week after the first dose of vaccination, in symptomatic individuals testing positive or negative for SARS-CoV-2. The colour bar represents the percentage of symptomatic individuals reporting each symptom.

Table 2 presents prevalence of each symptom over the week post-vaccination, divided into three windows. Some symptoms increased over time in both positive and negative individuals (e.g., headache, myalgia) whereas others increased in positive individuals only (e.g., sneezing, hoarse voice). Although fever and sore throat increased across the week in the negative individuals, there was a suggestion of a biphasic response in the positive individuals, also observed with persistent cough. The numbers of individuals were too small for formal testing; moreover, the exact date of infection in positive individuals was unknown.

**Table 2.** Symptom prevalence and timing during the first week post-vaccination (N=145), in symptomatic individuals testing positive or negative for SARS-CoV-2, grouped by days post-vaccination. Number of subjects and percentage (value in parenthesis).

|  | Positive SARS-CoV-2 |  |  | Negative SARS-CoV-2 |  |  |
| --- | --- | --- | --- | --- | --- | --- |
| Window | 0-2 days | 2-4 days | 4-7days | 0-2 days | 2-4 days | 4-7days |
| Headache | 22 (15.2) | 44 (30.3) | 66 (45.5) | 31 (21.4) | 46 (31.7) | 66 (45.5) |
| Chills or Shivers | 13 (9.0) | 25 (17.2) | 34 (23.4) | 21 (14.5) | 25 (17.2) | 26 (17.9) |
| Myalgias | 4 (2.8) | 12 (8.3) | 26 (17.9) | 7 (4.8) | 7 (4.8) | 20 (13.8) |
| Fever | 14 (9.7) | 10 (6.9) | 21 (14.5) | 11 (7.6) | 14 (9.7) | 20 (13.8) |
| Dizziness | 4 (2.8) | 13 (9.0) | 16 (11.0) | 6 (4.1) | 21 (14.5) | 20 (13.8) |
| Sneezing | 2 (1.4) | 8 (5.5) | 18 (12.4) | 1 (0.7) | 8 (5.5) | 6 (4.1) |
| Nausea | 5 (3.4) | 8 (5.5) | 11 (7.6) | 6 (4.1) | 15 (10.3) | 19 (13.1) |
| Sensitive Skin | 0 (0.0) | 6 (4.1) | 3 (2.1) | 0 (0.0) | 2 (1.4) | 2 (1.4) |
| Rhinorrhoea | 4 (2.8) | 10 (6.9) | 21 (14.5) | 1 (0.7) | 7 (4.8) | 19 (13.1) |
| Eye Soreness | 1 (0.7) | 6 (4.1) | 11 (7.6) | 4 (2.8) | 5 (3.4) | 8 (5.5) |
| Loss of Appetite | 6 (4.1) | 3 (2.1) | 19 (13.1) | 5 (3.4) | 6 (4.1) | 7 (4.8) |
| Chest Pain | 0 (0.0) | 3 (2.1) | 14 (9.7) | 1 (0.7) | 2 (1.4) | 5 (3.4) |
| Sore Throat | 9 (6.2) | 5 (3.4) | 30 (20.7) | 2 (1.4) | 6 (4.1) | 14 (9.7) |
| Brain Fog | 0 (0.0) | 4 (2.8) | 16 (11.0) | 5 (3.4) | 5 (3.4) | 10 (6.9) |
| Hoarse Voice | 1 (0.7) | 2 (1.4) | 15 (10.3) | 1 (0.7) | 2 (1.4) | 3 (2.1) |
| Anosmia | 0 (0.0) | 4 (2.8) | 2 (1.4) | 0 (0.0) | 2 (1.4) | 0 (0.0) |
| Abdominal Pain | 3 (2.1) | 3 (2.1) | 16 (11.0) | 2 (1.4) | 7 (4.8) | 15 (10.3) |
| Diarrhoea | 3 (2.1) | 4 (2.8) | 11 (7.6) | 2 (1.4) | 3 (2.1) | 9 (6.2) |
| Dysosmia | 1 (0.7) | 3 (2.1) | 12 (8.3) | 3 (2.1) | 1 (0.7) | 8 (5.5) |
| Ear Pain | 0 (0.0) | 3 (2.1) | 3 (2.1) | 1 (0.7) | 3 (2.1) | 4 (2.8) |
| Red Welts FL | 0 (0.0) | 1 (0.7) | 0 (0.0) | 0 (0.0) | 0 (0.0) | 1 (0.7) |
| Fatigue | 1 (0.7) | 3 (2.1) | 10 (6.9) | 6 (4.1) | 2 (1.4) | 2 (1.4) |
| Persistent Cough | 4 (2.8) | 1 (0.7) | 17 (11.7) | 0 (0.0) | 2 (1.4) | 3 (2.1) |
| Swollen Glands | 0 (0.0) | 1 (0.7) | 6 (4.1) | 3 (2.1) | 3 (2.1) | 1 (0.7) |
| Blisters on Feet | 0 (0.0) | 0 (0.0) | 4 (2.8) | 0 (0.0) | 0 (0.0) | 0 (0.0) |
| Dyspnoea | 0 (0.0) | 0 (0.0) | 0 (0.0) | 0 (0.0) | 0 (0.0) | 0 (0.0) |

Individual symptom duration is shown in Supplementary Table S8. There were no significant differences in symptom duration in positive vs. negative individuals after first vaccination. Importantly, symptom assessment was truncated at seven days, noting as above that some symptoms were increasing in prevalence with time. Interestingly, amongst individuals testing negative, dysosmia and delirium had the longest duration (median, 2 days for each).

Similar results for symptom prevalence after first vaccination were obtained comparing the positive population with the constructed cohort (1:1 matched) of negative individuals selected by bootstrapping (Supplementary Table S9) (Supplementary Figure S1); and with the negative population as a whole (Supplementary Figure S2). Some symptoms were significantly more common in negative individuals when using the entire negative population (e.g., brain fog), driven by the extremely large negative sample size, which supports our use of a selected matched population to avoid bias from unbalanced sample size.

There was no correlation between symptoms in either the positive or negative populations, assessed using Spearman-rank test (Supplementary Figure S3). As a sensitivity analysis we assessed the impact of a (self-logged) previous COVID-19 diagnosis; this made no difference to our results.

### SARS-CoV-2 test outcome prediction modelling

Model performance including receiver operator curves, using all reported symptoms, are shown in Table 3 and Figure 4. The best performance was obtained with random forest, followed by logistic regression; however, neither reached clinical utility. Other models, including clinical symptom clustering (Supplementary Table S4) and categorisation of individuals using NHS screening criteria, were no better than chance.

**Table 3.** Model performance in the classification of COVID-19 status according to post-vaccination symptoms. Median values and percentiles [0.25 and 0.75] are obtained across five folds. AUC - area under curve, in a receiver operating characteristic analysis.

|  | Sensitivity | Specificity | ROC - AUC |
| --- | --- | --- | --- |
| Bayesian Mixed-Effect Model | 0.52 [0.47; 0.56] | 0.55 [0.47; 0.60] | 0.52 [0.47; 0.56] |
| Logistic Regression | 0.63 [0.58; 0.67] | 0.67 [0.60; 0.72] | 0.62 [0.58; 0.67] |
| Random Forest | 0.61 [0.58; 0.64] | 0.63 [0.56; 0.69] | 0.66 [0.61; 0.70] |
| Symptom clustering | 0.51 [0.49; 0.56] | 0.67 [0.60; 0.73] | 0.50 [0.47; 0.55] |
| NHS screening criteria | 0.48 [0.48; 0.48] | 0.62 [0.61; 0.63] | 0.47 [0.47; 0.48] |

**Figure 4.**
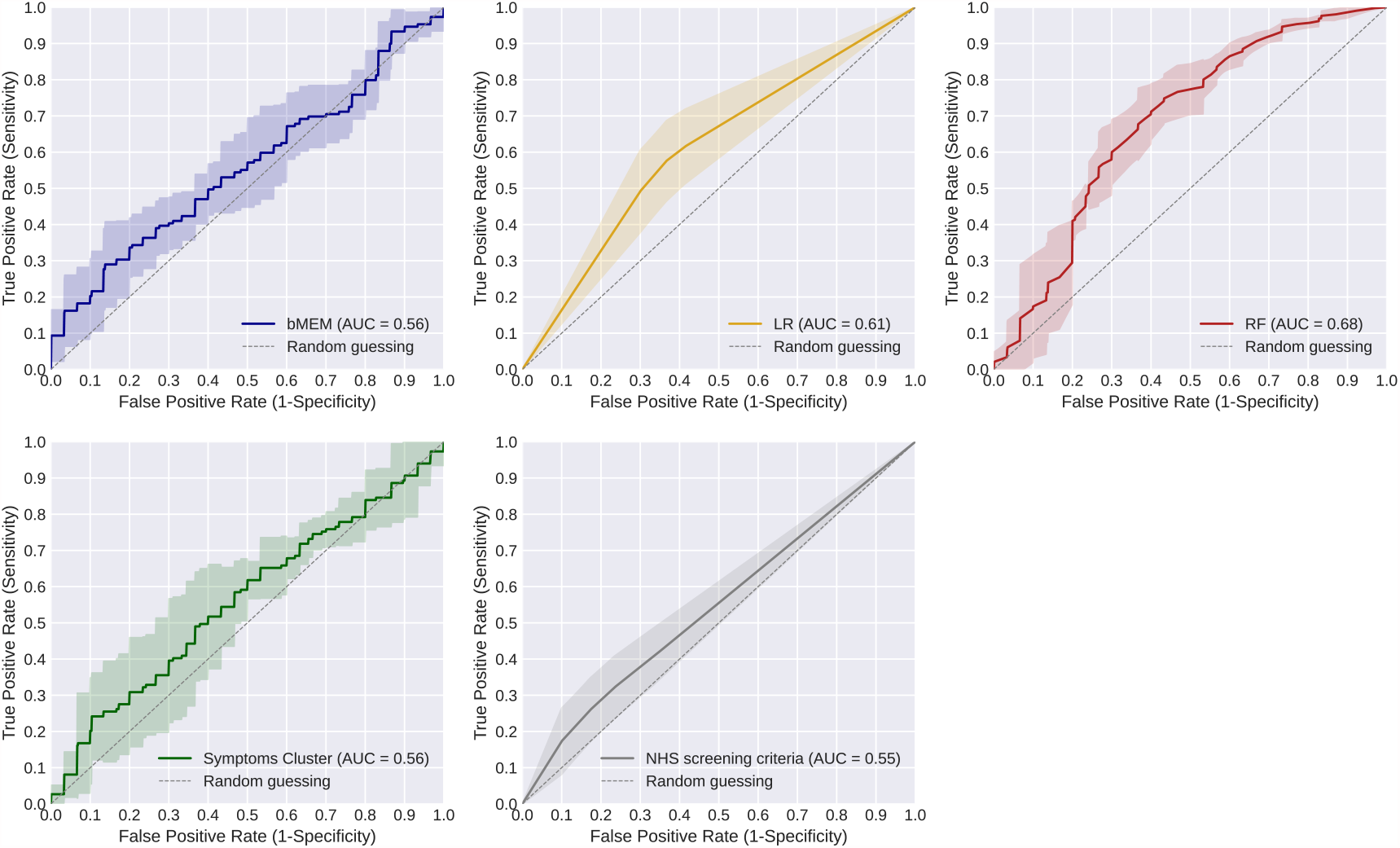
ROC-AUC performance for the different models. Mean value (line) and 95% CI (shadow area) of the models’ performance, given the predictions obtained over the cross-validation scheme (5-folds) adopted for the validation. bMEM: Bayesian Mixed-effect Model (blue); LR: Logistic Regression (yellow); RF: Random Forest (red); Symptoms clustering (green) and NHS screening criteria (grey).

## Discussion

Here we aimed to develop a clinically useful algorithm predictive of SARS-CoV-2 infection early post-vaccination, by parsing symptoms according to proven infection status in symptomatic individuals. Such an algorithm would be extremely useful, particularly in countries with limited health resources, as testing could be targeted towards those predicted positive, with quarantining of these individuals until an available result. To our knowledge, this is the first study with this aim. However, we were unable to differentiate post-vaccination symptoms *per se* from superimposed SARS-CoV-2 infection with robustness. Although two models showed ROC AUC significantly greater than 0.5, neither came close to approaching clinical utility - for most clinical tests, conventionally given as 0.8; but for a highly infectious agent with devastating consequences from community spread the necessary AUC is much higher.

Although one third of the one million vaccinated app users reported symptoms previously associated with COVID-19 early post-vaccination, only 4% of symptomatic individuals reported testing for SARS-CoV-2 even with allowance for delayed testing access. Considering those individuals who reported at least one of the symptoms fulfilling NHS criteria for testing (266,502 overall), 40% (107,929) were tested. During the study period, testing was widely available in the UK and it is unclear why more symptomatic people (including those with the widely advertised core symptoms) were not tested.^33^ Conversely, of 149 individuals who tested positive, only 62 (41%) had symptoms that met current UK testing criteria. We do not know why the other 88 positive individuals were tested (e.g., contact tracing, routine workplace testing, direct personal request through the app).

Our data also suggest sensitivity of using core symptoms for COVID-19 may be lower post-vaccination than in pre-vaccination times (here 48%, previously 73%.).^34^ Although individuals with core symptoms were more likely to test positive than those without, the overall sensitivity and AUC suggests current UK testing policy is suboptimal for pandemic management particularly now that rapid testing capacity is much greater than when these criteria were established. ^34^ Notably, current UK testing criteria are more limited than WHO guidelines^2^ and that of many other jurisdictions of similar GDP (including France, Germany, USA, and Australia).

Although there were some differences in symptom prevalence and distribution between positive and negative individuals, these could not be used robustly to discriminate between groups, including using machine-learning. We also considered time of symptom onset and symptom duration post-vaccination, (previous trials and post-marketing observational data have examined these parameters but not with respect to SARS-CoV-2 status).^4–6,8,16^ Whether positive or negative, median symptom peak burden was day 3 in both groups, concordant with vaccination side-effect profiles reported previously. ^4–6,8,16^ As time progressed, some symptoms started to become more common in the positive group only (e.g., persistent cough, hoarse voice), which timing coincides with the serial interval and incubation period of SARS-CoV-2;^26^ however, the critical public health importance of identifying and isolating cases early does not allow the luxury of a watch-and-wait approach.

We do not know the circumstances contributing to infection of the positive group (whether prior to, peri-, or immediately post-vaccination), noting here that we required rtPCR or LFAT results thus confirming recent infection. The serial interval and incubation period for SARS-CoV-2, and the high proportion of asymptomatic infection means that individuals could have been infected before vaccination. Although current UK vaccination guidelines do not require individuals to be completely asymptomatic at time of vaccination (only not “acutely unwell”^35^), our inclusion criteria required individuals to be asymptomatic at time of vaccination. It is also possible, that infection was contracted whilst getting vaccinated. Nosocomial infection with SARS-CoV-2 has been reported in the UK,^36,37^ including many health care workers infected at their workplace.^38,39^ Here we would emphasise strongly that data from ourselves and others indicate that even if infected peri- or post-vaccination, the course of COVID-19 is much less severe in vaccinated vs. unvaccinated individuals;^40–44^ and - acknowledging that a small percentage of vaccinated and symptomatic individuals were tested - our data demonstrated only 150 cases of confirmed infection (1%) in 14,842 tested individuals from over 1 million vaccinated app users. It is also possible that individuals changed their behaviour immediately after vaccination, increasing infection risk and contracting SARS-CoV-2 prior to acquiring adequate immunity. Here, data from previous infections suggest vaccination reduces adherence to other public health measures,^45^ with pre-prints suggesting that this is occurring after vaccination against SARS-CoV-2 also.^46,47^

Overall, CSS app users are not fully representative of the UK population (younger, more likely to be female, of higher educational status, and over-representative of healthcare workers)^21^. Although the population in the current study shares some of these biases, the median age of vaccinated individuals at the time of our analysis (64 years) was older than for app users overall (47 years), which is not surprising as the UK vaccination schedule began with the oldest individuals in the community. We considered the implications of this with respect to the likelihood of an infected person presenting for testing: although asymptomatic SARS-CoV-2 infection is well-recognised, it is less common in older people.^25,26^ We also acknowledge that different economic and cultural experiences may influence presentation of SARS-CoV-2 infection and reporting of post-vaccination side-effects.^45^

Our approach in comparing symptom profiles for individuals testing positive or negative for SARS-CoV-2 required a 1:1 matched population, so that comparison of symptom prevalence was fair and unbiased by the greatly different sample sizes of the two populations. However, this methodological choice is less reliable when used for the outcome of SARS-CoV-2 test prediction; and the forced balance of the classes can lead to an overestimation of the likelihood of being positive in the modelling. Thus we have also presented extended analyses, using both boot-strapping, and entire-cohort approaches in the Supplementary Results. We acknowledge that the predictive power of our optimised models may be hampered if there are brand-specific post-vaccination side-effects, which were not considered during model optimisation.^16^ However, although there were some differences in frequencies, most symptoms were reported in both PB and O-AZ pivotal trials.^4,5,8,10^ We also did not consider type of SARS-CoV-testing (PCR vs. LFAT), or mode of testing access (NHS vs. ZOE-request), which may also contribute variability to these models.

Our analyses do not consider the impact of COVID-19 prevalence in UK at the time of the vaccination. Positive and negative predictive values (PPV, NPV) for a test depend not only on test sensitivity and specificity but also on population prevalence of disease. The rapidly changing prevalence of SARS-CoV-2 infection in the UK and the pace of vaccination delivery over the time period of this study limits our capacity to provide accurate PPV and NPV. Further analyses, particularly in populations with higher prevalence of infection and/or higher symptom burden and severity of COVID-19, may result in better differentiation of early signs of infection from post-vaccination side-effects.

A strength of our study was our very large cohort of vaccinated participants, in a country that was an early adopter of vaccination; and our timeframe included the UK pandemic “third wave”. Prospective real-time symptom logging through the app minimised recall bias; and our symptom assessment included direct ascertainment of core symptoms for accessing UK testing. However, the sharp decline in cases in the first six months of 2021 resulted in only 150 positive cases to inform our modelling which number we acknowledge is small - though we were also able to draw upon large numbers of tested negative individuals for comparisons, reinforcing the consistency and generalisation of our results. We also acknowledge that the demographic features of the app population especially those parameters considered for model estimation (e.g., age, gender, BMI) may be different in other populations within the UK and elsewhere.

The implication of our results will vary depending on the population prevalence of SARS-CoV-2 and pace of vaccination roll-out. At the time of writing Australia has negligible community spread of SARS-CoV-2 but is early in vaccination roll-out. It would be very interesting to repeat this study in these different circumstances. Testing of all symptomatic individuals comes at a cost (e.g., testing kits, infrastructure). Here, the UK is a resource-rich country; the impact of our results in countries with fewer health resources needs careful consideration.

In conclusion, post-vaccination symptoms cannot be distinguished with clinical confidence from early SARS-CoV-2 infection. Our study highlights the critical importance of testing symptomatic individuals - even if recently vaccinated – to ensure early detection of SARS-CoV-2 infection and help prevent future waves of COVID-19.

## Data Availability

Data collected in the COVID Symptom Study smartphone application are shared with other health researchers through the UK National Health Service-funded Health Data Research UK (HDRUK) and Secure Anonymised Information Linkage consortium, housed in the UK Secure Research Platform (Swansea, UK). Anonymised data are available to be shared with researchers according to their protocols in the public interest (https://web.www.healthdatagateway.org/dataset/fddcb382-3051-4394-8436-b92295f14259). The code is available in: https://gitlab.com/KCL-BMEIS/covid-zoe/vaccination.

https://web.www.healthdatagateway.org/dataset/fddcb382-3051-4394-8436-b92295f1425

## Acknowledgements

ZOE Limited provided in-kind support for all aspects of building, running and supporting the app and service to all users worldwide. This work is supported by the Wellcome EPSRC Centre for Medical Engineering at King’s College London (WT 203148/Z/16/Z) and the UK Department of Health via the National Institute for Health Research (NIHR) comprehensive Biomedical Research Centre award to Guy’s & St Thomas’ NHS Foundation Trust in partnership with King’s College London and King’s College Hospital NHS Foundation Trust. Investigators also received support from Medical Research Council (MRC), British Heart Foundation (BHF), Alzheimer’s Society, European Union, NIHR, COVID-19 Driver Relief Fund (CDRF) and the NIHR-funded BioResource, Clinical Research Facility and Biomedical Research Centre (BRC) based at GSTT NHS Foundation Trust in partnership with KCL. SO was supported by the French government, through the 3IA Côte d’Azur Investments in the Future project managed by the National Research Agency (ANR) with the reference number ANR-19-P3IA-0002. This research was funded in part by the Wellcome Trust [215010/Z/18/Z]. For the purpose of Open Access, the author has applied a CC BY public copyright licence to any Author Accepted Manuscript (AAM) version arising from this submission.

## Authors’ contribution

LSC, MM and ELD contributed to study concept and design. CHS, JCP, BM, TS, CJS, SO contributed to acquisition of data. All the authors had access to the raw data underlying the study. LSC, JD, ELD contributed to data analysis and verified the underlying data. LSC, MO, MM and ELD contributed to drafting of the manuscript. All authors contributed to interpretation of data and critical revision of the manuscript. MM and ELD contributed to study supervision.

## Declaration of Interests

ELD and CJS report grants from the Chronic Disease Research Foundation (CDRF) during the conduct of the study. CH, SS, LP, AM report other from ZOE Limited, during the conduct of the study. TS is a scientific advisor to ZOE Limited. CHS reports grants from Alzheimer’s Society, during the conduct of the study. SO reports grants from the Wellcome Trust, Innovate UK (UKRI), and Chronic Disease Research Foundation (CDRF), during the conduct of the study.

## Abbreviations

AUC: Area under the curve
bMEM: Bayesian mixed-effect model
BMI: Body mass index
CI: confidence interval
COVID-19: Coronavirus disease 2019
CSS: COVID Symptom Study
IQR: inter-quartile range
KCL: King’s College London
LFAT: Lateral flow antigen test
LR: Logistic regression
MD: Missing data
NHS UK: National Health Service of the United Kingdom
O-AZ: Oxford-AstraZeneca adenovirus-vectored vaccine
PB: Pfizer-BioNTech mRNA vaccine
RF: Random forest
ROC: Receiver operating curve
rtPCR: Reverse transcription polymerase chain reaction
SARS-CoV-2: severe acute respiratory syndrome-related coronavirus 2
UK: United Kingdom of Great Britain and Northern Ireland

## Notes

### Author Declarations

The app and CSS were approved in the UK by KCL's ethics committee (REMAS no. 18210, review reference LRS 19/20 18210). All app users provided informed consent for use of their data for COVID-19 research.

