## Supplementary material for "Disentangling post-vaccination symptoms from early COVID-19"

### Supplementary Materials

Contents:

- Supplementary Methods
    - App development
    - Testing Kits
- Supplementary Results
    - Tables:
        - Supplementary Table S1. List of symptom questions asked by the COVID Symptom Study app.
        - Supplementary Table S2. List of local symptoms caused by the COVID Symptom Study app, caused by vaccination.
        - Supplementary Table S3: Properties and features of models used in this study.
        - Supplementary Table S4. Listing of clinical symptoms grouping.
        - Supplementary Table S5. Profiles of illness in symptomatic individuals after the 1st dose of vaccination (N=145) and 2nd dose (N=4).
        - Supplementary Table S6. Symptom prevalence and distribution during the first week after vaccination, in symptomatic individuals testing positive for SARS-CoV-2 infection.
        - Supplementary Table S7. Symptom prevalence and distribution during the first week after vaccination, in symptomatic individuals testing negative for SARS-CoV-2 infection (1:1 matched cohort).
        - Supplementary Table S8. Duration of individual symptoms after first vaccination (irrespective of symptom prevalence) in individuals testing positive or negative for SARS-CoV-2 (N=145 for each cohort).
        - Supplementary Table S9. Demographic information of vaccinated individuals testing positive (N=149) and a constructed cohort of equal size created by bootstrapping testing negative for SARS-CoV-2.
    - Figures:
        - Supplementary Figure S1. Profiles of illness in symptomatic individuals early post-vaccination, comparing symptom prevalence (symptom reported at any time during first week) in positive vs. negative cases (N=145 for each cohort), using bootstrapping to construct the negative cohort.
        - Supplementary Figure S2. Profiles of illness in symptomatic individuals early post-vaccination, comparing symptom prevalence (symptom reported at any time during first week) in individuals testing positive (N=145) vs. negative (N=12112) for SARS-CoV-2 infection.
        - Supplementary Figure S3. Correlation of symptoms experienced early post-vaccination in individuals testing positive (left image) or negative (right image) for SARS-CoV-2 infection (N=149).

### Supplementary Methods

#### App development

In this prospective cohort study, data were acquired from CSS, through a mobile application for iPhone® and Android® users launched jointly by ZOE Limited. and KCL on 24 March 2020. [15] Software was tested before app launch, and before release of any version update; software for repeatable and consistent data extraction, curation, and analytics was also engineered. [48]

#### Testing Kits

SARS-CoV-2 testing kits were supplied by the Department of Health and Social Care, UK Government. These were sent to app users on their request.

### Supplementary Results: Tables

**Supplementary Table S 1. List of symptom questions asked by the COVID Symptom Study app.** Questions were: Do you have (symptom)? Answers were yes/no, unless indicated otherwise.

| Symptom | COVID Symptom Study app question |
|---|---|
| Fever | Fever (at least 37.8C or 100F) |
| Persistent Cough | Persistent cough (coughing a lot for more than an hour or 3 or more coughing episodes in 24 hours) |
| Fatigue | Unusual fatigue… (no; mild fatigue; severe fatigue/ I struggle to get out of bed) |
| Dyspnoea | Shortness of breath or trouble breathing (no; yes mild symptoms/ slight shortness of breath during ordinary activity: yes significant symptoms/ breathing is comfortable only at rest; yes, severe symptoms/ breathing is difficult even at rest). |
| Anosmia/Ageusia | Loss of smell / taste |
| Hoarse Voice | Unusually hoarse voice |
| Chest Pain | Unusual chest pain or tightness in your chest |
| Abdominal Pain | Unusual abdominal pain or stomach ache |
| Diarrhoea | Diarrhoea |
| Delirium | Confusion, disorientation or drowsiness |
| Eye Soreness (ophthalmalgia) | Do your eyes have any unusual eye-soreness or discomfort (e.g. light sensitivity, excessive tears, or pink/red eye)? |
| Loss of appetite (anorexia) | Skipping meals |
| Headache | Headache |
| Nausea | Nausea or vomiting |
| Dizziness | Dizziness or light-headedness |
| Sore Throat | Sore or painful throat |
| Myalgias | Unusual strong muscle pains or aches |
| Red Welts in Face and Lips | Raised, red, itchy welts on the skin or sudden swelling of the face or lips |
| Blisters on feet | Red/purple sores or blisters on your feet, including your toes |
| Sensitive Skin | Strange, unpleasant sensations in your skin like pins & needles or burning |
| Brain Fog | Loss of concentration or memory (brain fog) |
| Dysosmia/Dysgeusia | Altered smell / taste (things smell or taste different to usual) |
| Rhinorrhoea | Runny nose |
| Sneezing (sternutation) | Sneezing more than usual |
| Ear Pain (otalgia) | Earache |

| | |
|---|---|
| Lymphadenopathy | Swollen neck glands |
| Palpitations | Irregular heart beat |

**Supplementary Table S 2. List of local symptoms caused by the COVID Symptom Study app, caused by vaccination. Questions were: Do you have (symptom)? Answers were yes/no, unless indicated otherwise.** These symptoms were excluded from this analysis.

| Symptom | COVID Symptom Study app question |
|---|---|
| Tenderness | Tenderness near the site of injection |
| Pain | Arm pain |
| Redness | Redness near the site of injection |
| Swelling | Arm or local swelling |
| Warmth | Warmth near the site of injection |
| Bruising | Bruising near the site of injection |
| Swollen glands | Swollen glands in the armpit |

**Supplementary Table S 3. Properties and features of models used in this study.**

| Models | Input variables | Response variable | Model Characteristics | COVID-19 studies |
|---|---|---|---|---|
| Bayesian mixed-effect model | Fixed effects: all symptoms;<br><br>Random effects: demographic information (age, BMI, gender) | Test outcome – probabilistic outcome | Parametric model [49] | N/A |
| Logistic regression | All symptoms, and demographic information | Test outcome – binary outcome | Parametric model | COVID-19 detection using self-reported symptoms [21] |
| Random forest | All symptoms, and demographic information | Test outcome – binary outcome | Non-parametric model - intrinsic feature space transformation | Identification of early stage symptoms of SARS-Cov-2 infected patients [50] |
| Clinical symptom clustering | All symptoms | Test outcome | | N/A |
| NHS diagnostic criteria | Fever, Cough, Anosmia, Dysosmia | Test outcome | Screening criteria adopted by NHS | COVID-19 test referencing |

**Supplementary Table S 4. Listing of clinical symptoms grouping.**

| Clinical grouping | List of symptoms |
|---|---|
| Upper respiratory tract symptoms | anosmia, sore throat, rhinorrhoea, sneezing, ear pain, dysosmia |
| Respiratory | persistent cough, dyspnoea, chest pain, hoarse voice, plus any/all of the upper respiratory tract list |
| Systemic | fever, fatigue, delirium, headache, chills or shivers, eye soreness, myalgias, dizzy, light-headed, swollen glands [other than axillary lymph nodes in vaccinated arm], brain fog, irregular heartbeat |
| Abdominal | diarrhoea, abdominal pain, nausea, loss of appetite |
| Cutaneous | blisters on feet, sensitive skin and red welts on face and lips |

**Supplementary Table S 5. Profiles of illness in symptomatic individuals after the 1st dose of vaccination (N=145) and 2nd dose (N=4).** Cells show absolute number (percentage in parentheses) of symptomatic individuals for symptoms reported at any time during first week, in positive vs. negative cases**.**

| | First dose (N=145) | | Second dose (N=4) | |
|---|---|---|---|---|
| **Symptoms** | **Positive test for SARS-CoV-2 infection** | **Negative test for SARS-CoV-2 infection** | **Positive test for SARS-CoV-2 infection** | **Negative test for SARS-CoV-2 test infection** |
| Headache | 86 (59.3) | 101 (70.0) | 3 (0.75) | 3 (0.75) |
| Chills or Shivers | 56 (38.6) | 60 (41.3) | 0 (0.0) | 1(0.25) |
| Fever | 40 (27.6) | 36 (24.8) | 1(0.25) | 0 (0.0) |
| Sore Throat | 32 (22.1) | 117 (11.7) | 1(0.25) | 0 (0.0) |
| Myalgias | 29 (20.0) | 25 (17.2) | 2 (0.50) | 0 (0.0) |
| Rhinorrhoea | 27 (18.6) | 19 (13.1) | 2 (0.50) | 1(0.25) |
| Sneezing | 23 (15.9) | 12 (8.3) | 1(0.25) | 0 (0.0) |
| Nausea | 22 (15.2) | 26 (17.9) | 0 (0.0) | 0 (0.0) |
| Dizziness | 22 (15.2) | 31 (21.4) | 0 (0.0) | 1(0.25) |
| Loss of Appetite | 19 (13.1) | 17 (11.7) | 0 (0.0) | 0 (0.0) |
| Abdominal Pain | 16 (11.0) | 18 (12.4) | 0 (0.0) | 2 (0.50) |
| Diarrhoea | 14 (9.7) | 10 (6.9) | 0 (0.0) | 0 (0.0) |
| Eye Soreness | 12 (8.3) | 13 (9.0) | 0 (0.0) | 0 (0.0) |

| | | | | |
|---|---|---|---|---|
| Persistent Cough | 12 (8.3) | 4 (2.8) | 0 (0.0) | 0 (0.0) |
| Hoarse Voice | 12 (8.3) | 5 (3.4) | 2 (0.50) | 0 (0.0) |
| Chest Pain | 11 (7.6) | 7 (4.8) | 0 (0.0) | 0 (0.0) |
| Delirium | 10 (6.9) | 5 (3.4) | 0 (0.0) | 0 (0.0) |
| Fatigue | 9 (6.2) | 9 (6.2) | 0 (0.0) | 1(0.25) |
| Brain Fog | 9 (6.2) | 13 (9.0) | 0 (0.0) | 0 (0.0) |
| Dysosmia | 7 (4.8) | 6 (4.1) | 1(0.25) | 0 (0.0) |
| Sensitive Skin | 6 (4.1) | 4 (2.8) | 0 (0.0) | 0 (0.0) |
| Anosmia | 5 (3.4) | 2 (1.4) | 0 (0.0) | 0 (0.0) |
| Ear Pain | 4 (2.8) | 6 (4.1) | 0 (0.0) | 0 (0.0) |
| Swollen Glands | 4 (2.8) | 6 (4.1) | 1(0.25) | 0 (0.0) |
| Palpitations | 3 (2.1) | 11 (7.6) | 0 (0.0) | 0 (0.0) |
| Red Welts FL | 1(0.7) | 1(0.7) | 0 (0.0) | 0 (0.0) |
| Blisters on Feet | 1(0.7) | 0 (0.0) | 0 (0.0) | 0 (0.0) |
| Dyspnoea | 0 (0.0) | 0 (0.0) | 0 (0.0) | 0 (0.0) |

**Supplementary Table S 6. Symptom prevalence and distribution during the first week after vaccination, in symptomatic individuals testing positive for SARS-CoV-2 infection.** Data report number (percentage in parentheses) of individuals with each symptom on each day after first vaccination (N = 145).

| | **Positive SARS-CoV-2** | | | | | | | |
|---|---|---|---|---|---|---|---|---|
| **Days after vaccination** | 0 | 1 | 2 | 3 | 4 | 5 | 6 | 7 |
| Headache | 1 (0.7) | 10 (6.9) | 11 (7.6) | 29 (20.0) | 15 (10.3) | 12 (8.3) | 23 (15.9) | 31 (21.4) |
| Chills or Shivers | 0 (0.0) | 6 (4.1) | 7 (4.8) | 15 (10.3) | 10 (6.9) | 10 (6.9) | 12 (8.3) | 12 (8.3) |
| Myalgias | 1 (0.7) | 1 (0.7) | 2 (1.4) | 10 (6.9) | 2 (1.4) | 3 (2.1) | 8 (5.5) | 15 (10.3) |
| Fever | 0 (0.0) | 8 (5.5) | 6 (4.1) | 8 (5.5) | 2 (1.4) | 5 (3.4) | 8 (5.5) | 8 (5.5) |
| Dizziness | 0 (0.0) | 2 (1.4) | 2 (1.4) | 7 (4.8) | 6 (4.1) | 3 (2.1) | 5 (3.4) | 8 (5.5) |
| Sneezing | 0 (0.0) | 1 (0.7) | 1 (0.7) | 6 (4.1) | 2 (1.4) | 4 (2.8) | 2 (1.4) | 12 (8.3) |
| Nausea | 0 (0.0) | 3 (2.1) | 2 (1.4) | 6 (4.1) | 2 (1.4) | 1 (0.7) | 3 (2.1) | 7 (4.8) |
| Sensitive Skin | 0 (0.0) | 0 (0.0) | 0 (0.0) | 6 (4.1) | 0 (0.0) | 0 (0.0) | 3 (2.1) | 0 (0.0) |
| Rhinorrhoea | 0 (0.0) | 3 (2.1) | 1 (0.7) | 4 (2.8) | 6 (4.1) | 3 (2.1) | 7 (4.8) | 11 (7.6) |
| Eye Soreness | 0 (0.0) | 1 (0.7) | 0 (0.0) | 4 (2.8) | 2 (1.4) | 0 (0.0) | 4 (2.8) | 7 (4.8) |
| Loss of Appetite | 0 (0.0) | 4 (2.8) | 2 (1.4) | 3 (2.1) | 0 (0.0) | 4 (2.8) | 7 (4.8) | 8 (5.5) |
| Chest Pain | 0 (0.0) | 0 (0.0) | 0 (0.0) | 3 (2.1) | 0 (0.0) | 1 (0.7) | 10 (6.9) | 3 (2.1) |
| Sore Throat | 1 (0.7) | 3 (2.1) | 5 (3.4) | 3 (2.1) | 2 (1.4) | 2 (1.4) | 14 (9.7) | 14 (9.7) |
| Brain Fog | 0 (0.0) | 0 (0.0) | 0 (0.0) | 3 (2.1) | 1 (0.7) | 2 (1.4) | 7 (4.8) | 7 (4.8) |
| Hoarse Voice | 0 (0.0) | 1 (0.7) | 0 (0.0) | 2 (1.4) | 0 (0.0) | 1 (0.7) | 5 (3.4) | 9 (6.2) |
| Anosmia | 0 (0.0) | 0 (0.0) | 0 (0.0) | 2 (1.4) | 2 (1.4) | 0 (0.0) | 1 (0.7) | 1 (0.7) |
| Abdominal Pain | 0 (0.0) | 2 (1.4) | 1 (0.7) | 2 (1.4) | 1 (0.7) | 6 (4.1) | 4 (2.8) | 6 (4.1) |
| Diarrhoea | 0 (0.0) | 1 (0.7) | 2 (1.4) | 2 (1.4) | 2 (1.4) | 2 (1.4) | 0 (0.0) | 9 (6.2) |
| Dysosmia | 0 (0.0) | 1 (0.7) | 0 (0.0) | 2 (1.4) | 1 (0.7) | 0 (0.0) | 7 (4.8) | 5 (3.4) |
| Ear Pain | 0 (0.0) | 0 (0.0) | 0 (0.0) | 2 (1.4) | 1 (0.7) | 0 (0.0) | 0 (0.0) | 3 (2.1) |
| Red Welts FL | 0 (0.0) | 0 (0.0) | 0 (0.0) | 1 (0.7) | 0 (0.0) | 0 (0.0) | 0 (0.0) | 0 (0.0) |
| Fatigue | 0 (0.0) | 1 (0.7) | 0 (0.0) | 1 (0.7) | 2 (1.4) | 0 (0.0) | 4 (2.8) | 6 (4.1) |
| Persistent Cough | 0 (0.0) | 0 (0.0) | 4 (2.8) | 0 (0.0) | 1 (0.7) | 4 (2.8) | 7 (4.8) | 6 (4.1) |
| Swollen Glands | 0 (0.0) | 0 (0.0) | 0 (0.0) | 0 (0.0) | 1 (0.7) | 0 (0.0) | 4 (2.8) | 2 (1.4) |
| Blisters on Feet | 0 (0.0) | 0 (0.0) | 0 (0.0) | 0 (0.0) | 0 (0.0) | 0 (0.0) | 4 (2.8) | 0 (0.0) |
| Dyspnoea | 0 (0.0) | 0 (0.0) | 0 (0.0) | 0 (0.0) | 0 (0.0) | 0 (0.0) | 0 (0.0) | 0 (0.0) |

**Supplementary Table S 7. Symptom prevalence and distribution during the first week after vaccination, in symptomatic individuals testing negative for SARS-CoV-2 infection (1:1 matched cohort).** Data report number (percentage in parentheses) of individuals with each symptom on each day after first vaccination (N=145).

| | **Negative SARS-CoV-2** | | | | | | | |
|---|---|---|---|---|---|---|---|---|
| **Days after vaccination** | 0 (0.0) | 1 (0.7) | 2 | 3 | 4 | 5 | 6 | 7 |
| Headache | 1 (0.7) | 13 (9.0) | 17 (11.7) | 16 (11.0) | 30 (20.7) | 23 (15.9) | 17 (11.7) | 26 (17.9) |
| Chills or Shivers | 0 (0.0) | 11 (7.6) | 10 (6.9) | 8 (5.5) | 17 (11.7) | 7 (4.8) | 5 (3.4) | 14 (9.7) |
| Myalgias | 0 (0.0) | 2 (1.4) | 5 (3.4) | 1 (0.7) | 6 (4.1) | 8 (5.5) | 5 (3.4) | 7 (4.8) |
| Fever | 0 (0.0) | 6 (4.1) | 5 (3.4) | 6 (4.1) | 8 (5.5) | 7 (4.8) | 7 (4.8) | 6 (4.1) |
| Dizziness | 0 (0.0) | 4 (2.8) | 2 (1.4) | 11 (7.6) | 10 (6.9) | 8 (5.5) | 7 (4.8) | 5 (3.4) |
| Sneezing | 0 (0.0) | 1 (0.7) | 0 (0.0) | 4 (2.8) | 4 (2.8) | 1 (0.7) | 1 (0.7) | 4 (2.8) |
| Nausea | 1 (0.7) | 2 (1.4) | 3 (2.1) | 4 (2.8) | 11 (7.6) | 3 (2.1) | 3 (2.1) | 13 (9.0) |
| Sensitive Skin | 0 (0.0) | 0 (0.0) | 0 (0.0) | 1 (0.7) | 1 (0.7) | 0 (0.0) | 0 (0.0) | 2 (1.4) |
| Rhinorrhoea | 0 (0.0) | 0 (0.0) | 1 (0.7) | 2 (1.4) | 5 (3.4) | 11 (7.6) | 2 (1.4) | 6 (4.1) |
| Eye Soreness | 0 (0.0) | 2 (1.4) | 2 (1.4) | 4 (2.8) | 1 (0.7) | 5 (3.4) | 3 (2.1) | 0 (0.0) |
| Loss of Appetite | 0 (0.0) | 0 (0.0) | 5 (3.4) | 2 (1.4) | 4 (2.8) | 3 (2.1) | 2 (1.4) | 2 (1.4) |
| Chest Pain | 0 (0.0) | 0 (0.0) | 1 (0.7) | 1 (0.7) | 1 (0.7) | 2 (1.4) | 2 (1.4) | 1 (0.7) |
| Sore Throat | 0 (0.0) | 1 (0.7) | 1 (0.7) | 2 (1.4) | 4 (2.8) | 9 (6.2) | 2 (1.4) | 3 (2.1) |
| Brain Fog | 0 (0.0) | 0 (0.0) | 5 (3.4) | 4 (2.8) | 1 (0.7) | 1 (0.7) | 6 (4.1) | 3 (2.1) |
| Hoarse Voice | 0 (0.0) | 0 (0.0) | 1 (0.7) | 0 (0.0) | 2 (1.4) | 0 (0.0) | 1 (0.7) | 2 (1.4) |
| Anosmia | 0 (0.0) | 0 (0.0) | 0 (0.0) | 1 (0.7) | 1 (0.7) | 0 (0.0) | 0 (0.0) | 0 (0.0) |
| Abdominal Pain | 0 (0.0) | 1 (0.7) | 1 (0.7) | 3 (2.1) | 4 (2.8) | 4 (2.8) | 7 (4.8) | 4 (2.8) |
| Diarrhoea | 0 (0.0) | 0 (0.0) | 2 (1.4) | 0 (0.0) | 3 (2.1) | 5 (3.4) | 1 (0.7) | 3 (2.1) |
| Dysosmia | 0 (0.0) | 1 (0.7) | 2 (1.4) | 0 (0.0) | 1 (0.7) | 4 (2.8) | 0 (0.0) | 4 (2.8) |
| Ear Pain | 0 (0.0) | 1 (0.7) | 0 (0.0) | 1 (0.7) | 2 (1.4) | 0 (0.0) | 1 (0.7) | 3 (2.1) |
| Red Welts FL | 0 (0.0) | 0 (0.0) | 0 (0.0) | 0 (0.0) | 0 (0.0) | 0 (0.0) | 0 (0.0) | 1 (0.7) |
| Fatigue | 0 (0.0) | 2 (1.4) | 4 (2.8) | 2 (1.4) | 0 (0.0) | 0 (0.0) | 1 (0.7) | 1 (0.7) |
| Persistent Cough | 0 (0.0) | 0 (0.0) | 0 (0.0) | 0 (0.0) | 2 (1.4) | 3 (2.1) | 0 (0.0) | 0 (0.0) |
| Swollen Glands | 0 (0.0) | 2 (1.4) | 1 (0.7) | 1 (0.7) | 2 (1.4) | 0 (0.0) | 1 (0.7) | 0 (0.0) |
| Blisters on Feet | 0 (0.0) | 0 (0.0) | 0 (0.0) | 0 (0.0) | 0 (0.0) | 0 (0.0) | 0 (0.0) | 0 (0.0) |
| Dyspnoea | 0 (0.0) | 0 (0.0) | 0 (0.0) | 0 (0.0) | 0 (0.0) | 0 (0.0) | 0 (0.0) | 0 (0.0) |

**Supplementary Table S 8. Duration of individual symptoms after first vaccination (irrespective of symptom prevalence) in individuals testing positive or negative for SARS-CoV-2 (N=145 for each cohort).** Median value of symptom duration ([0.25; 0.75] quantiles) is detailed per symptom. Differences in duration distribution per symptom were assessed using Mann-Whitney-U tests. P-value refers to the result of the statistical test.

| Symptoms | Positive SARS-CoV-2 | Negative SARS-CoV-2 | P-Value |
|---|---|---|---|
| Headache | 1 [1; 2] | 1 [1; 2] | 0.4428 |
| Chills or Shivers | 1 [1; 1] | 1 [1; 1] | 0.7550 |
| Myalgias | 1 [1; 2] | 1 [1; 1] | 0.4489 |
| Fever | 1 [1; 1] | 1 [1; 1] | 0.1599 |
| Dizziness | 1 [1; 2] | 1 [1; 2] | 0.7303 |
| Sneezing | 1 [1; 1] | 1 [1; 1.25] | 0.4415 |
| Nausea | 1 [1; 1] | 1 [1; 1.75] | 0.1033 |
| Sensitive Skin | 1 [1; 1.75] | 1 [1; 1] | 0.6667 |
| Rhinorrhoea | 1 [1; 1] | 1 [1; 1] | 0.9383 |
| Eye Soreness | 1 [1; 2] | 1 [1; 1] | 0.3634 |
| Loss of Appetite | 1 [1; 1.5] | 1 [1; 1] | 0.1015 |
| Chest Pain | 1 [1; 2] | 1 [1; 1] | 0.4314 |
| Sore Throat | 1 [1; 2] | 1 [1; 1] | 0.1747 |
| Brain Fog | 2 [1; 3] | 1 [1; 2] | 0.2129 |
| Hoarse Voice | 1 [1; 1.25] | 1 [1; 1] | 1.0000 |
| Anosmia | 1 [1; 1] | 1 [1; 1] | 1.0000 |
| Abdominal Pain | 1 [1; 1.25] | 1 [1; 1] | 0.8707 |
| Diarrhoea | 1 [1; 1] | 1 [1; 2] | 0.0896 |
| Dysosmia | 1 [1; 3.5] | 2 [1; 2] | 0.9825 |
| Ear Pain | 1 [1; 1.5] | 1 [1; 1.75] | 1.0000 |
| Red Welts FL | 1 [1; 1] | 1 [1; 1] | 1.0000 |
| Fatigue | 1 [1; 1] | 1 [1; 1] | 0.7353 |
| Persistent Cough | 1.5 [1; 2.25] | 1 [1; 1.25] | 0.5077 |
| Swollen Glands | 1 [1; 1.75] | 1 [1; 1] | 0.8000 |
| Blisters on Feet | 4 [4; 4] | N/A | N/A |
| Dyspnoea | N/A | N/A | N/A |
| Delirium | 1 [1; 1.75] | 2 [1; 3] | 0.4775 |
| Palpitations | 1 [1; 1] | 1 [1; 1.5] | 0.9066 |

**Supplementary Table S 9. Demographic information of vaccinated individuals testing positive (N=149) and a constructed cohort of equal size created by bootstrapping testing negative for SARS-CoV-2.** Demographic information for the negative SARS-CoV-2 cohort was estimated based on the selected bootstrapped samples: thus, for most values the range is presented (as [Minimum; Maximum]); when appropriate median value [IQR] is presented (age, BMI). BMI: Body Mass Index. IQR: Inter Quartile Range.

| | **Vaccinated Cohort** | | | | | |
|---|---|---|---|---|---|---|
| | **Positive test for SARS-CoV-2 infection** | | | **Negative test for SARS-CoV-2 infection** | | |
| | **O-AZ** | **PB** | **Full cohort** | **O-AZ** | **PB** | **Full cohort** |
| Number | 72 | 77 | 149 | 72 | 77 | 149 |
| Males (%) | 37.0 | 27.6 | 32.2 | [36.9; 37.0] | [27.6; 27.6] | [32.2; 32.2] |
| Age, years (median [IQR]) | 62.0 [50.0; 71.0] | 59.0 [50.0; 67.5] | 61.0 [50.0; 70.0] | 62.1 [49.3; 71.0] | 59.4 [50.5; 67.8] | 60.7 [50.3; 70.4] |
| BMI (median [IQR]) | 25.0 [22.7; 28.0] | 26.1 [23.5; 29.3] | 25.4 [23.4; 29.2] | 25.1 [22.9; 28.3] | 26.1 [23.6; 29.5] | 25.6 [23.2; 29.0] |
| Lung disease (%) | 11.1 | 11.8 | 11.4 | [5.5; 15.1] | [5.3; 22.4] | [7.4; 14.1] |
| Kidney Disease (%) | 0.0 | 1.3 | 0.7 | [0.0; 4.1] | [0.0; 5.3] | [0.0; 3.4] |
| Diabetes (%) | 5.5 | 6.6 | 6.0 | [2.7; 10.9] | [2.6; 7.9] | [2.7; 8.1] |
| Heart Disease (%) | 9.7 | 5.3 | 7.4 | [2.7; 10.9] | [5.3; 11.8] | [4.0; 7.4] |
| Cancer (%) | 0.0 | 5.3 | 2.7 | [0.0; 6.9] | [0.0; 3.9] | [0.7; 4.7] |
| Healthcare workers (%) | 0.0 | 13.2 | 6.7 | [0.0; 2.7] | [6.6; 14.5] | [0.7; 5.4] |
| Visit to hospital (%) | 1.4 | 1.3 | 1.3 | [0.0; 1.4] | [0.0; 3.9] | [0.0; 2.0] |

### Supplementary Results: Figures

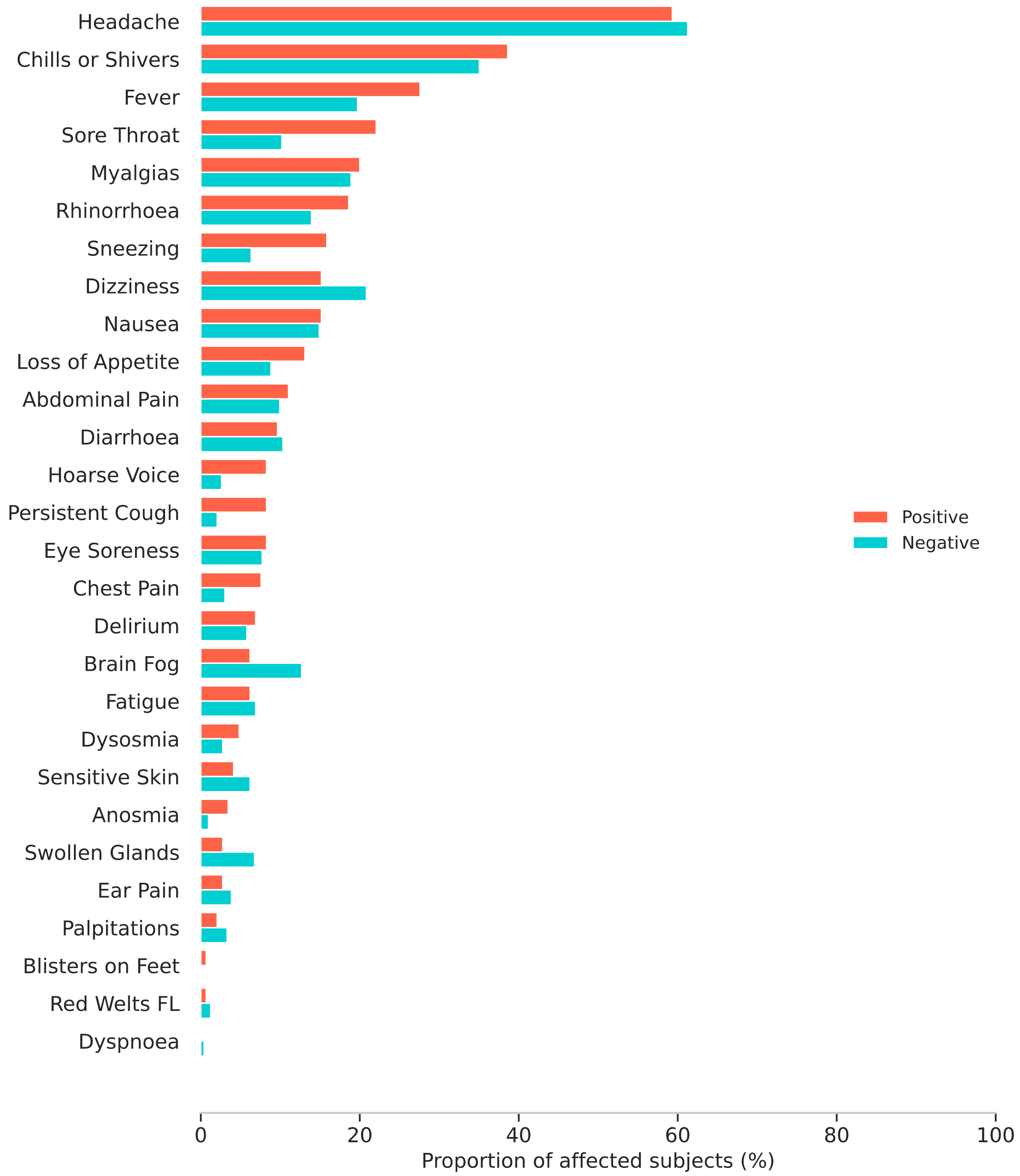


**Supplementary Figure S 1. Profiles of illness in symptomatic individuals early post-vaccination, comparing symptom prevalence (symptom reported at any time during first week) in positive vs. negative cases (N=145 for each cohort), using bootstrapping to construct the negative cohort.** The confidence interval (black bar) for the negative population is due to use of bootstrapping to create the negative population.

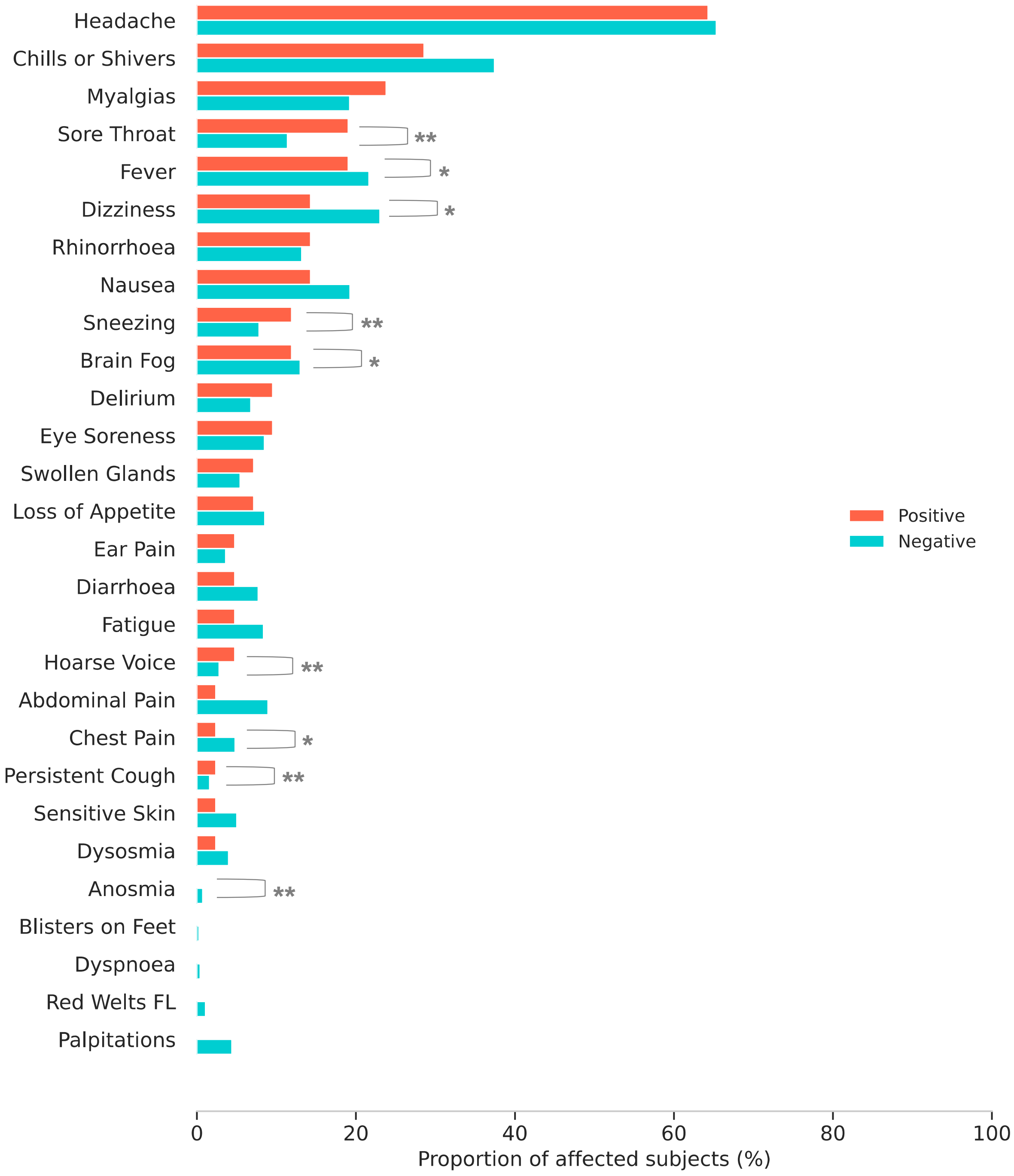


**Supplementary Figure S 2. Profiles of illness in symptomatic individuals early post-vaccination, comparing symptom prevalence (symptom reported at any time during first week) in individuals testing positive (N=145) vs. negative (N=12112) for SARS-CoV-2 infection.** *p < 0.05 **p < 0.01.

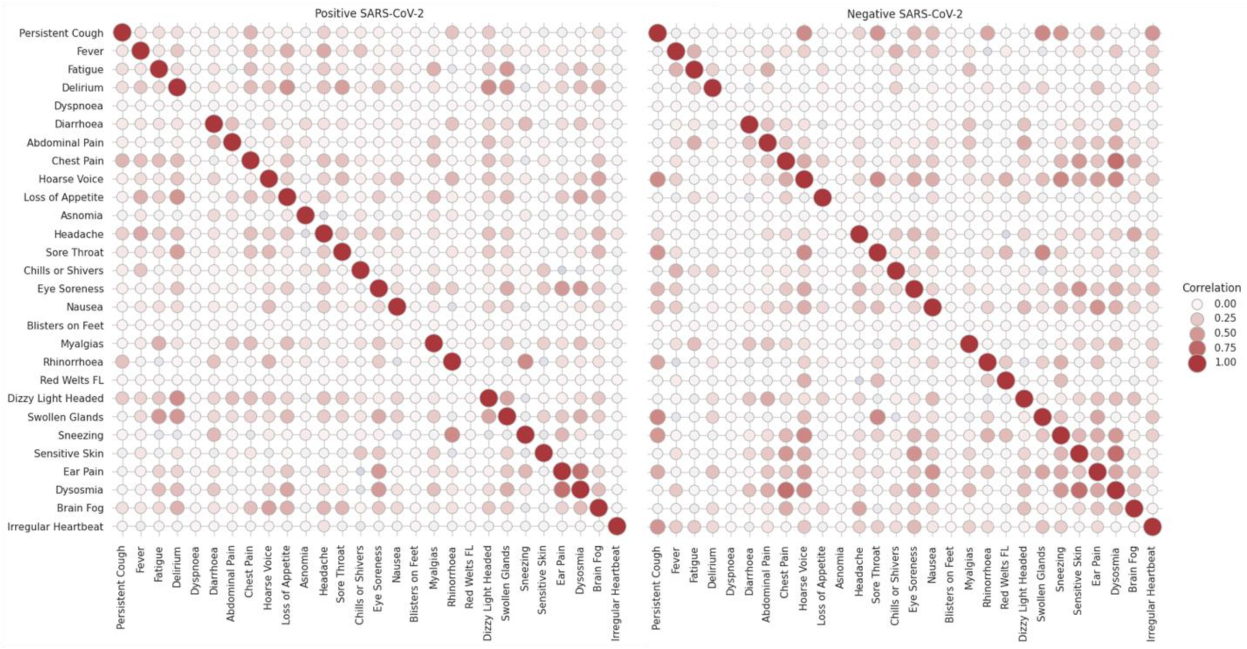


**Supplementary Figure S 3. Correlation of symptoms experienced early post-vaccination in individuals testing positive (left image) or negative (right image) for SARS-CoV-2 infection (N=149).** The colour and size of the marker encode the Spearman-rank correlation. Darker colours represent higher correlation of symptoms.
